# Estimating Prevalence and time Course of Sars-Cov-2 Based on new Hospital Admissions and PCR Tests: Relevance to Vaccination Program Tactical Planning

**DOI:** 10.1101/2020.08.15.20175653

**Authors:** Jose E. Gonzalez

## Abstract

Data posted in the COVID 19 tracking website for RT-PCR (PCR) results and hospital admissions are used to estimate the time course of the SARS-CoV-2 pandemic in the United States (1) and individual states. Hospital admissions mitigate positive sampling bias in PCR tests since these were limited in numbers initially. Additionally, their intent was as a diagnostic rather than a surveying tool.

By September 17, the United States’ cumulative recovered population is estimated at 45% or 149 million. The remaining susceptible population is 55%, or 50%, excepting the currently infected 5% population. The estimated mortality rate of the cumulative of the total affected population is 0.13% death.

States have followed diverse epidemic time courses. New Jersey and New York show SARS-CoV-2 prevalence of 95% and 82%, respectively. Likewise, each state exhibits relatively low current positive PCR results at 1.2 % and 0.8%. Also, these states show about twice the mortality rate of the nation. By comparison, Florida, California, and Texas showed recovered populations percent around 50%, and higher current PCR positive test results ranging from 5% to 9%.

This novel approach provides an improved source of information on the pandemic’s full-time course in terms of precision and accuracy in contrast to serological testing, which only views a narrow time slice of its history due to the transient nature of the antibody response and its graduated expression dependency on the severity of the disease. The deficiency of serological testing to estimate the recovered population is made even more acute due to the large proportion of asymptomatic and sub-clinical cases in the COVID-19 pandemic (2,3). T-cell testing, reputedly capable of long-term detection of previously infected individuals, will provide a complete view of the recovered population when it becomes available for large scale use.

This New Hospital Admission based method informs a more effective and efficient deployment of a vaccination program since it provides not only a reliable estimate of the susceptible population by state, but it can also provide visibility down to the county level based on COVID-19 hospitalization record independent of PCR testing.

## INTRODUCTION

The expectation to use simple statistics based on serological testing to measure the historical prevalence of SARS-CoV-2 was dashed upon discovering the transient and disease-severity indexed antibody response in COVID-19 patients (2,3). In the United States, 45% of the population (149 million people) have been infected by SARS-CoV-2 as of September 17. Longitudinal antibody monitoring data available from the states of Colorado, Georgia, South Carolina, and Texas fail to show, in the face of sustained infection rates over several months, the expected cumulative increase in the population carrying antibodies to SARS-CoV-2.

Recent T-cell tests suggest that infected individuals who appear negative in antibody tests have SARS-CoV-2 reactive T-cells. As many as 40% to 60% of blood samples collected before the start of the SARS-CoV-2 epidemic show B or T cells that cross-react with the virus (4, 5, 6, 7, 8). Taken together with the extensive level of infection cited earlier, it points ideally to deploying a vaccination campaign strategy that tests prospectively for SARS-CoV-2 reactive T or B cells in subjects before vaccination to make this treatment available to the more vulnerable first. Absent this assay tool, the quantitative estimation presented here allows for a more effective allocation of the vaccine to states with the larger susceptible populations. Moreover, this estimation can be applied to the county level, provided COVID-19 hospitalization records are available.

This paper relies on a novel approach that integrates % PCR positive test results over time, cycle-corrected for the length of disease, and coupled with hospital admissions to control for PCR testing sample bias, to estimate the kinetics and prevalence over the time course of the pandemic in the United States and individual states (3).

Early on the SARS-CoV-2 pandemic, studies relied on mortality rate, on the CDC reported influenza-like-illness (ILI) surge to explore prevalence (9), and serology to measure the extent of infection in New York state (10,11). By comparison, this paper bases its approach on PCR test records and COVID-19 specific hospital admissions posted by the COVID-19 Tracking Project (1). New hospital admissions correlate with the ILI surge reported in 2020. As with ILI, new hospital admissions are exempt from positive sampling bias.

Not only does the correlation with new hospital admissions enables the mitigation of positive sampling bias found in PCR test results due to its use as a diagnostic tool instead of as a population survey measure, but it also compensates for the early limited numbers of daily PCR tests performed, and the consequential high positive results detected. When PCR tests are plentiful, such that % positives fall to roughly 5% of the number of tests, and the ratio to new hospital admissions becomes stable, the PCR sampling bias is largely mitigated. The ratio obtained by dividing NHA by %PCR new cases in this stable region, in turn, enables the estimation of % positive cases in the population by dividing the observed NHA by this constant ratio on any date over the time course of the pandemic.

Additionally, it helps correct inflated % positive PCR results obtained more recently in individual states when these reduce their sampling numbers.

## RESULTS

Because PCR tests were rolled out with an emphasis as a diagnostic rather than a survey tool, and only sick people were encouraged to test, positive results are likely to be enhanced by sampling bias. As sampling numbers increase well beyond the number of sick people tested, the positive bias diminishes. Figure 1 shows that beyond 500,000 daily tests, the % PCR positive remains steady at around 7% instead of decreasing with increasing daily test numbers, as is expected from the resulting dilution of positive bias. Thus, positive sampling bias is mitigated above this daily test number.

**Figure 1.**
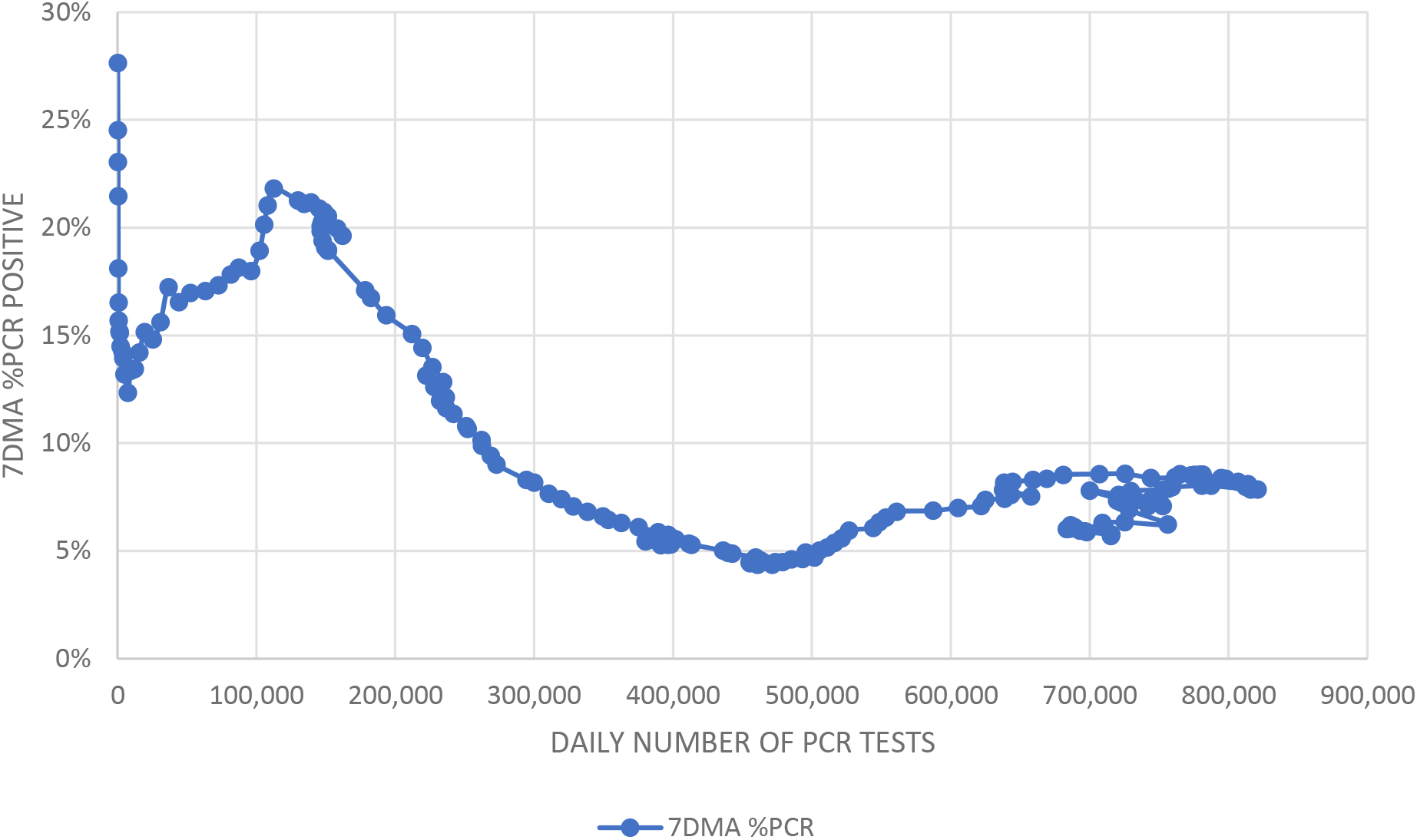
% RELATIONSHIP OF %PCR POSITIVES WITH TOTAL DAILY TEST RESULTS. Figure 1 shows the relationship of the 7-day moving average of positive % PCR versus the daily number of tests with the 7-day moving average of the total number of daily PCR tests. The % positive PCR stabilizes beyond 500,000 daily tests. 7-day moving averages are used in these figures and estimations because of the strong weekly cyclicality of data reporting.

A similar analysis is presented in Figure 2, which shows that the mean of the Ratio of New Hospital Admission (NHA) to % PCR New Cases remains within a narrow range (95% confidence interval of 2%) (about 516 admissions/%New PCR cases) when above 500,000 daily PCR tests. This constant ratio enables the estimation of % New Cases throughout the time course of the data unencumbered by the positive sampling bias attributed to the PCR test.

**Figure 2.**
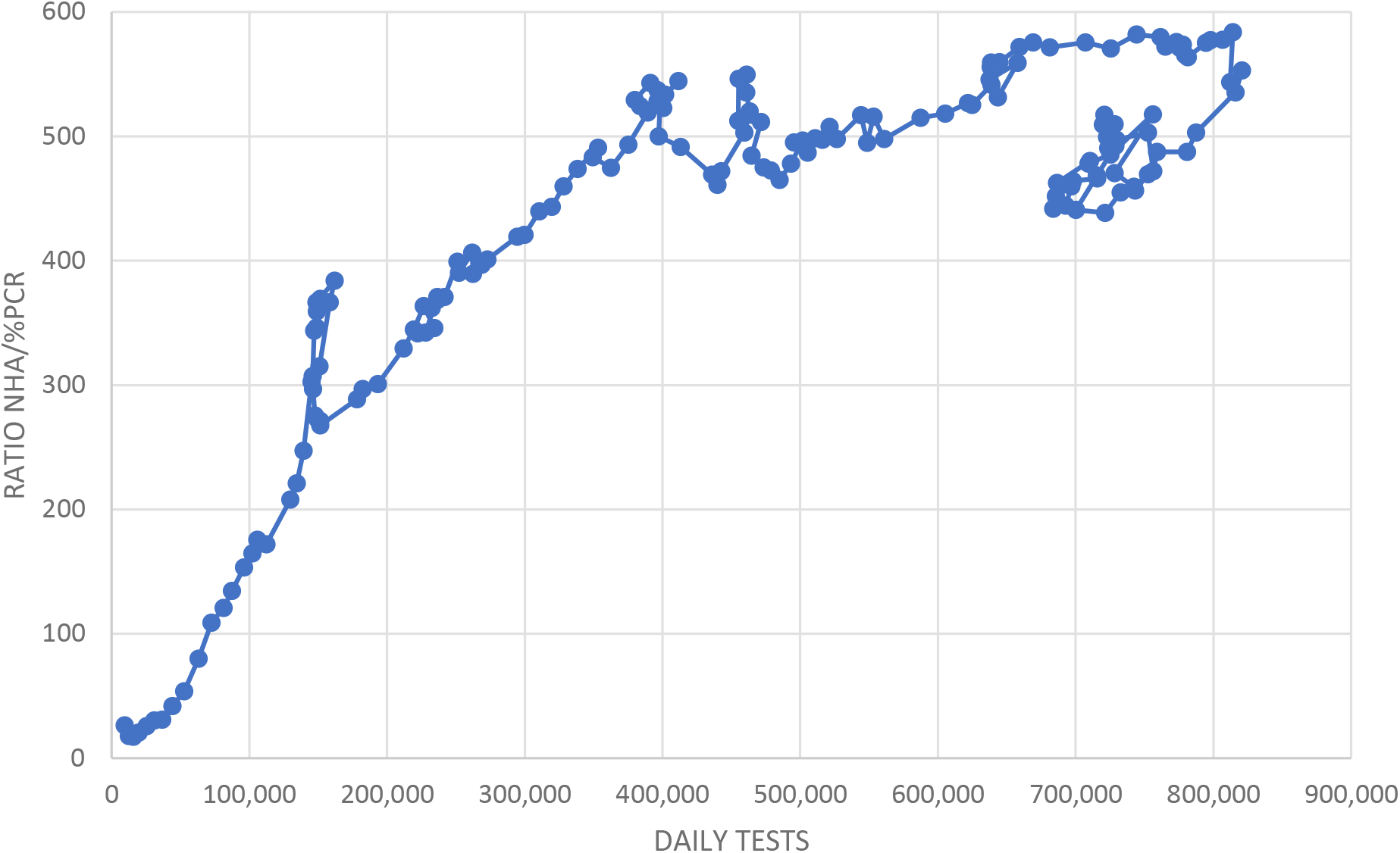
RELATIONSHIP OF NHA/%PCR RATIO WITH TOTAL TEST RESULTS. Figure 2 depicts the relationship between New Hospital Admissions (NHA) to %PCR positive ratio over increasing total PCR tests performed daily in the United States. Past 500,000 daily tests the ratio stabilizes about a value of 516. The graph is based on 7-day moving averages for NHA, %PCR positive, and test results. The 95% confidence interval of the mean is 10.5.

Figure 3A illustrates the positive bias correction obtained from the NHA curve, which shows lower values at the beginning of the pandemic (associated with lower daily test numbers) than % positive PCR tests. All curves are 7-day moving averages of the raw data reported by the COVID-19 Tracking Project (1). The NHA curve peaks at 14% while the %PCR curve draws a broad peak reaching 22% about April 11. The curves then trace converging paths to congruence on June 25, after which daily PCR tests consistently exceed 500,000.

**Figure 3A.**
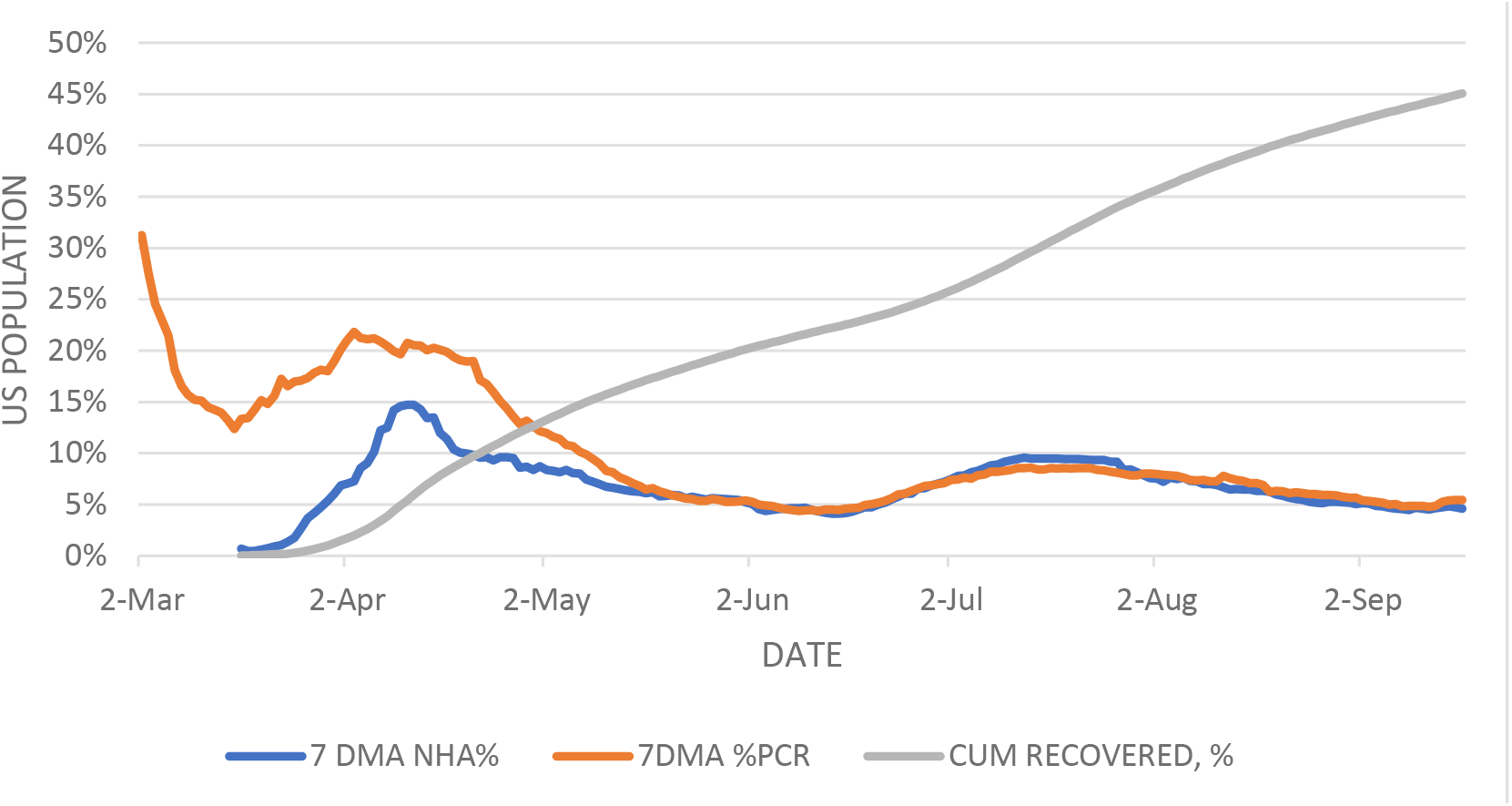
TIME COURSE COMPARISON OF PCR% CASES, NEW HOSPITAL ADMISSIONS DERIVED % CASES, AND RECOVERED % POPULATION. Figure 3A exhibits the relationship between the 7-day moving average of positive %PCR and NHA. After June 21, the daily number of PCR tests exceed 500,000. The ratio of these two variables is computed for the daily values after this date, obtaining an average of 516 through August 31 for the ratio of NHA/PCR % positive. The recovered population in the United States since March 17 is estimated here at 139 million.

Because the database (1) provides ‘currently hospitalized’ for 48 of the states (and ‘cumulative hospitalizations’ only for ten states), it becomes necessary to estimate the length of hospitalization to estimate NHA for all states (see Methods). The literature supports an average hospital stay of 12 days (12, 13, 14, 15, 16). Two states, Florida and Kansas, only report ‘cumulative hospitalized’ from which NHA is the difference between subsequent days. Of the remaining 48 states, ten reported both currently and cumulative hospitalizations, making it possible to compare the NHA estimating algorithm for both. This comparison confirms that the median hospital stay length of 12 days is the best fit to obtain concordance between the two reported datasets, cumulative versus currently hospitalized. Both medians and averages bracket the 12-day stay length reported in the literature, although the mean 95% confidence interval is about 25%.

Figure 3A also shows that the recovered population’s time course in the United States starting on March 17 reaches 45% by September 17. Since March 26, the number of infected people never falls below 12 million (3.6% of the U.S. population). Less than 1 million people were infected before March 17 (10, 11).

Estimating the cumulative recovered population from daily % positive cases requires knowing the time that PCR tests can detect the virus during the disease timeline. Literature estimates for the median time to conversion to PCR negative range from 17 to 28 days (14, 17, 18, 19, 20, 21). This paper uses the most extended time point (28 days) because it provides a lower boundary to the recovered population estimate.

In Figure 3B, the relationship with the CDC weekly P&I mortality prevalence is included. It peaks over the same period as %PCR cases and New Hospital Admissions (NHA). This agreement with P&I intensity reinforces NHA’s use as a marker for the prevalence of SARS-CoV-2 in the United States population. Normalizing the data for all three variables on week 21 shows a stable quantitative relationship between %PCR and NHA after that (R-square = 0.94), making it useful in estimating % positive cases of COVID-19 over the entire time based on NHA cases. The ILI incidence mortality report lags NHA and %PCR peaks by a couple of weeks in more recent dates.

**Figure 3B.**
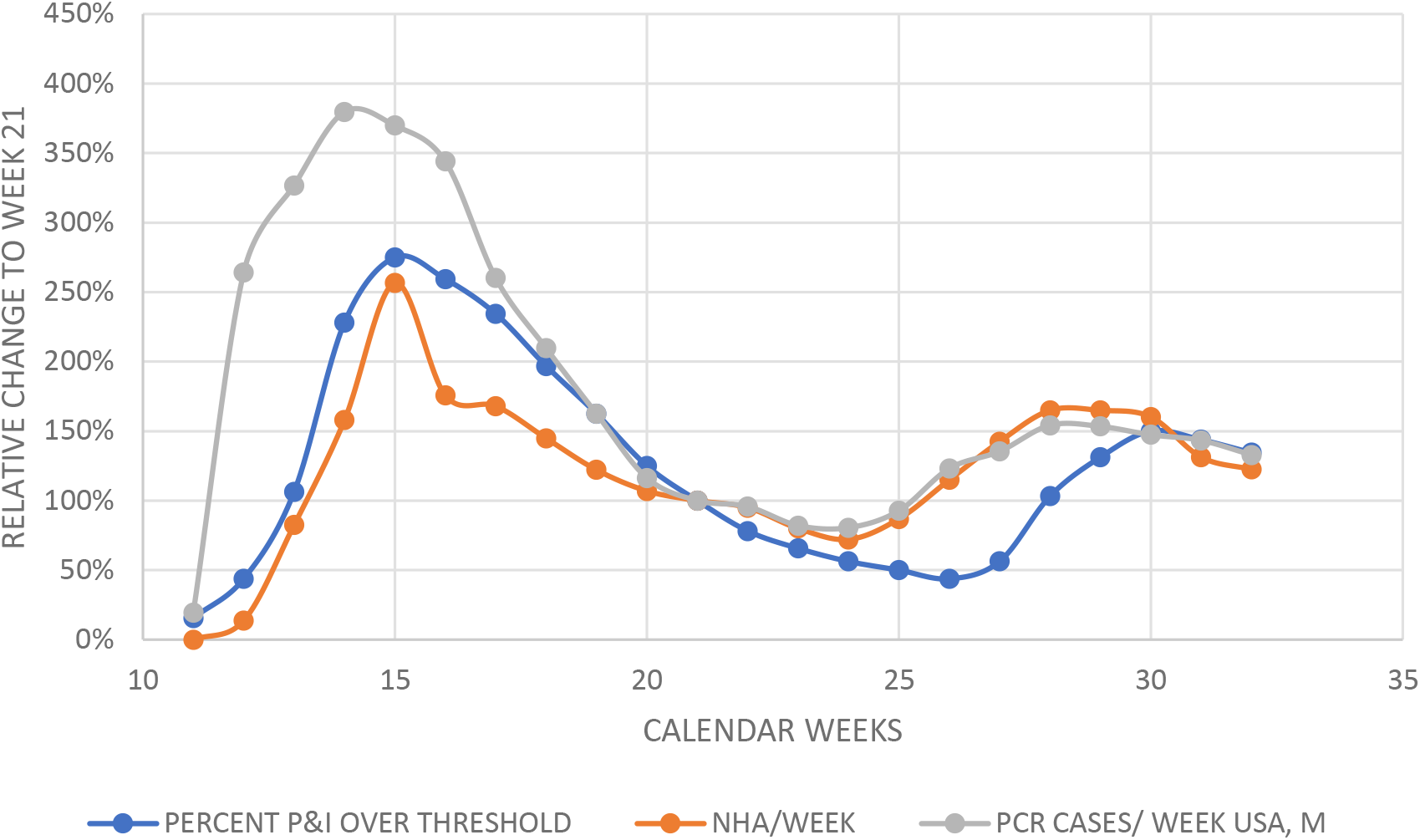
RELATIVE CHANGES OF CDC-ILI, NEW HOSPITAL ADMISSIONS, PCR POSITIVE CASES. Figure 3B shows the relationship of the CDC’s weekly report of P&I over the threshold, NHA, and New PCR cases to 100% on calendar week 21. All three variables tend to move together. NHA and PCR cases track tightly, starting on calendar week 21. Regression analysis of these two variables after that date yields a coefficient of determination of 0.94. As expected, percent P&I mortality over threshold tends to lag both cases by % PCR, and NHA derived cases.

The NHA derived infection time course curves allow the estimation of the number of people recovered nationwide. The daily curve values divided by the disease’s median detectable duration (28 days) give the total daily value of the recovered population in the United States (13, 14, 22). Cumulation of this value returns the total recovered population, depicted in Figure 5 in millions of people and population percent. Accordingly, by September 17, 45% of the United States population, or expressed in numbers, 149 million people had recovered from COVID-19, except for 189,665 deaths (1). Because on September 17, new cases in the United States registered at 4.6% (15 million people), the uninfected population remaining at risk is 50%, or 165 million. By comparison, the % PCR curve, with its positive early bias, results in an estimated 72% recovered population.

**Figure 4.**
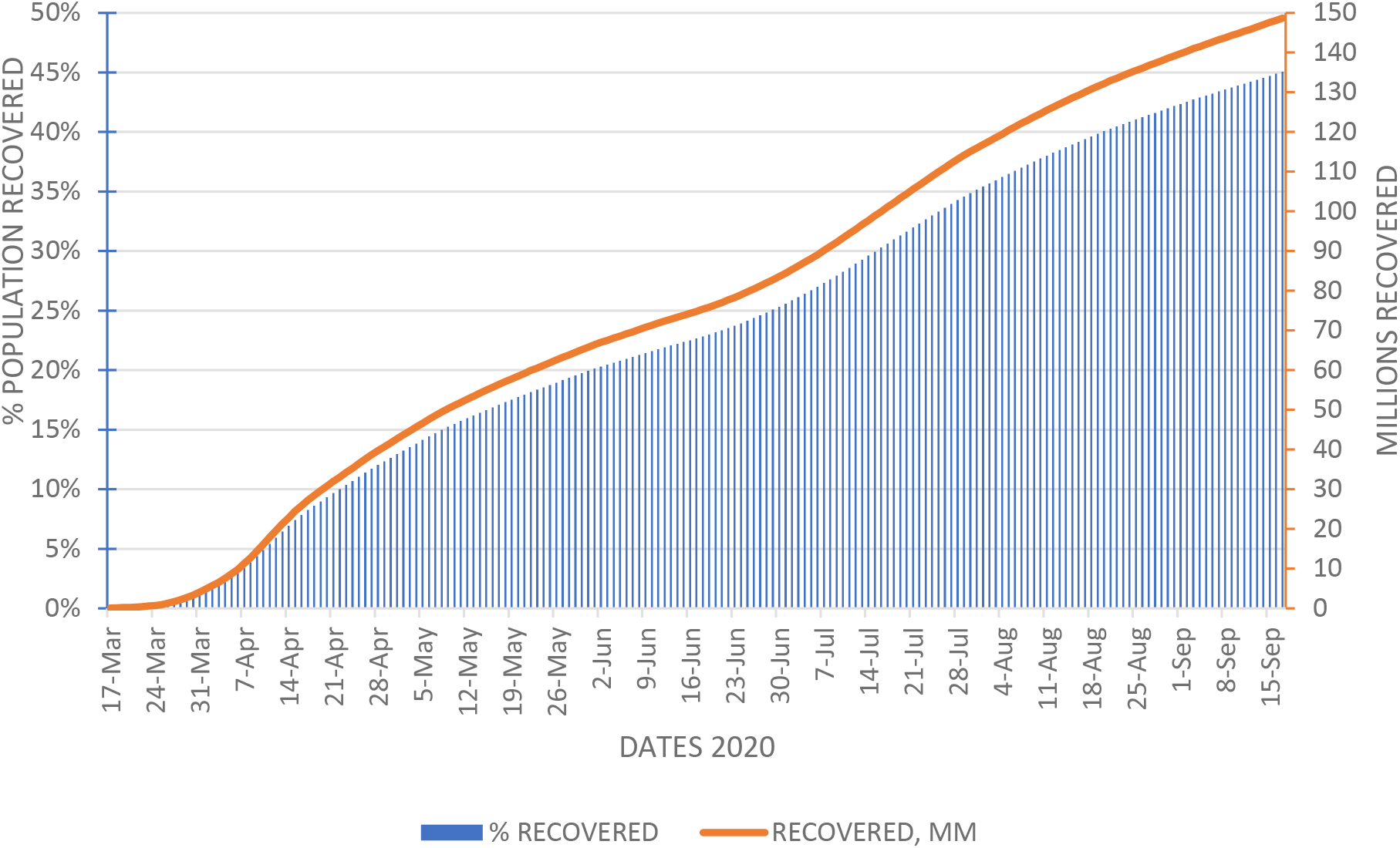
CUMULATIVE COVID-19 RECOVERED UNITED STATES POPULATION ON SEPTEMBER 17, 2020. Figure 4 illustrates the time course for the COVID-19 recovered population in terms of percent and millions of patients. This information is derived by summing the daily incidence obtained from the NHA new case curve divided by the duration of detectable disease (28 days) (13, 14, 22).

**Figure 5.**
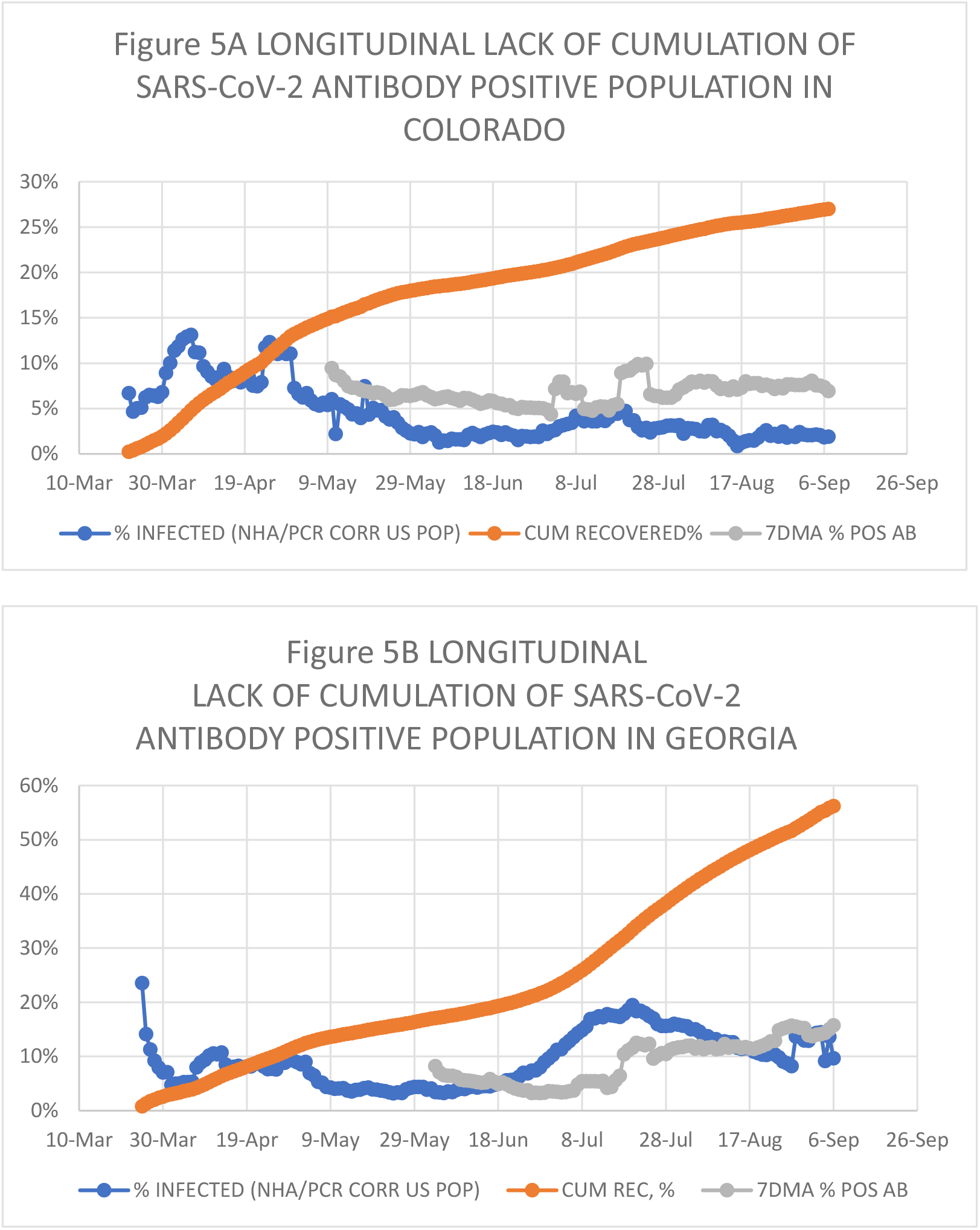
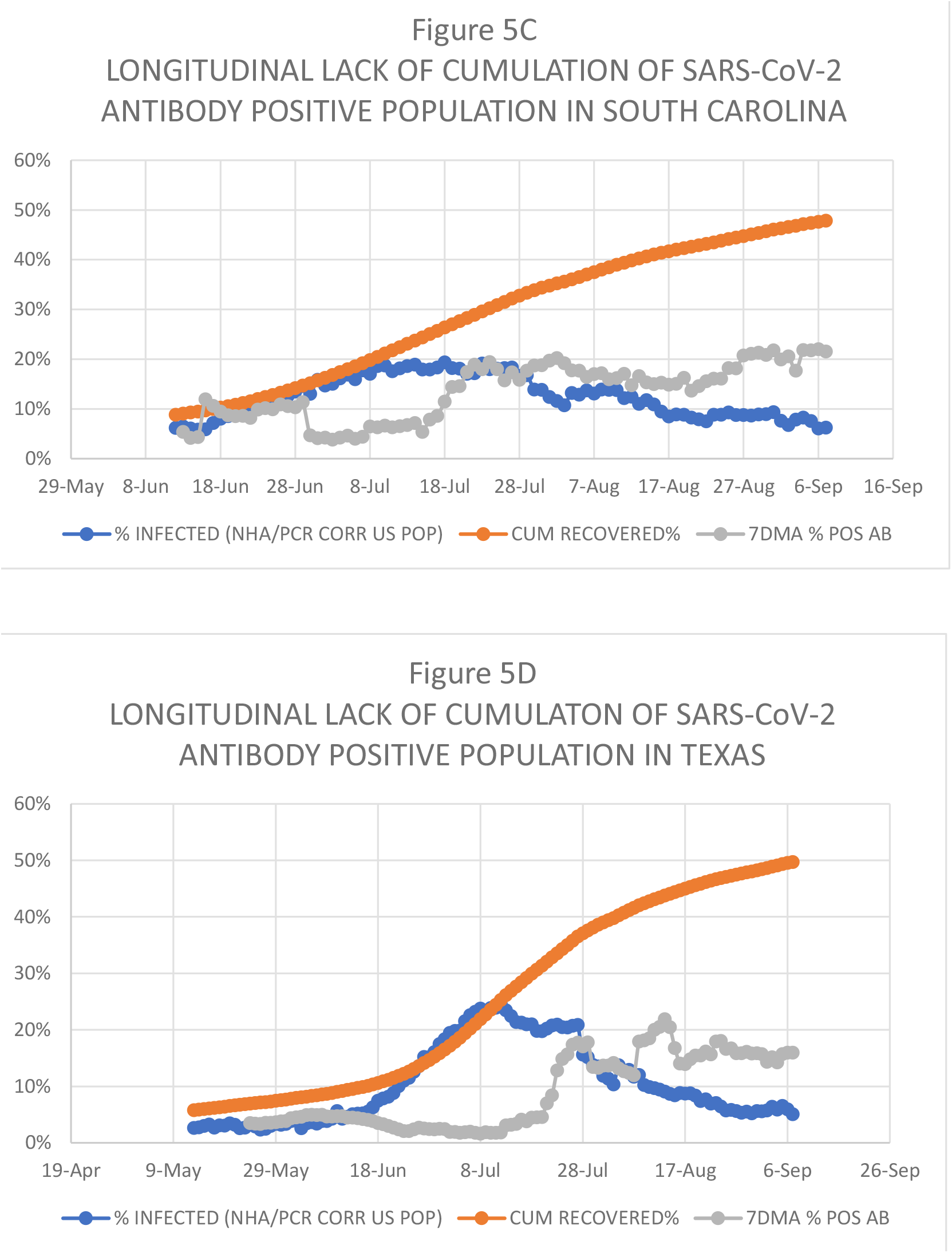
LONGITUDINAL STUDIES OF SARS-CoV-2 ANTIBODY CUMULATION IN THE POPULATION. Figure 5 shows the time course for the daily % infected population obtained from New Hospital Admissions (Blue), the Cumulative Recovered % population (Mustard), and Positive Antibody population % (Gray) for each of Colorado (a), Georgia (b), South Carolina (c), and Texas(d).

Table 1 summarizes the critical parameters of infection prevalence in the United States and individual states. Together with the densely populated and commercially connected Eastern seaboard states, the region from New Hampshire to the District of Columbia experienced a sudden and extensive onslaught of the epidemic in the Spring of 2020, which probably contributed to the high mortality rate in Massachusetts (0.33%), New Hampshire (0.25%), Connecticut (0.25%), Rhode Island (0.24%), Pennsylvania (0.22%), New York (0.21%), and New Jersey (0.19%), which compare poorly against the national average of 0.13%.

**Table 1.**
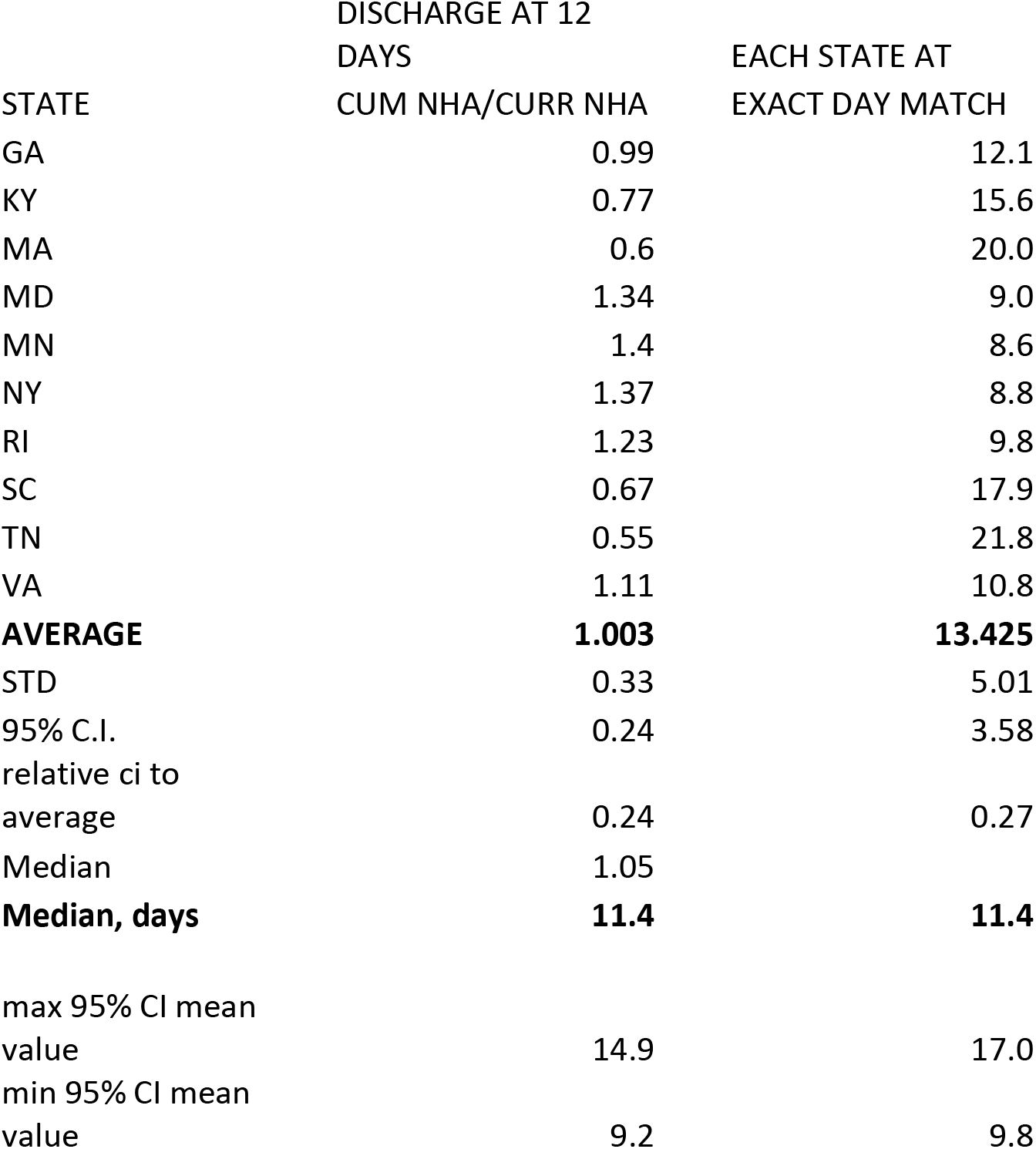
COMPARISON OF CUMULATIVE AND CURRENTLY HOSPITALIZED DATA TO COMPUTE NEW HOSPITAL ADMISSIONS.

**Table 2.**
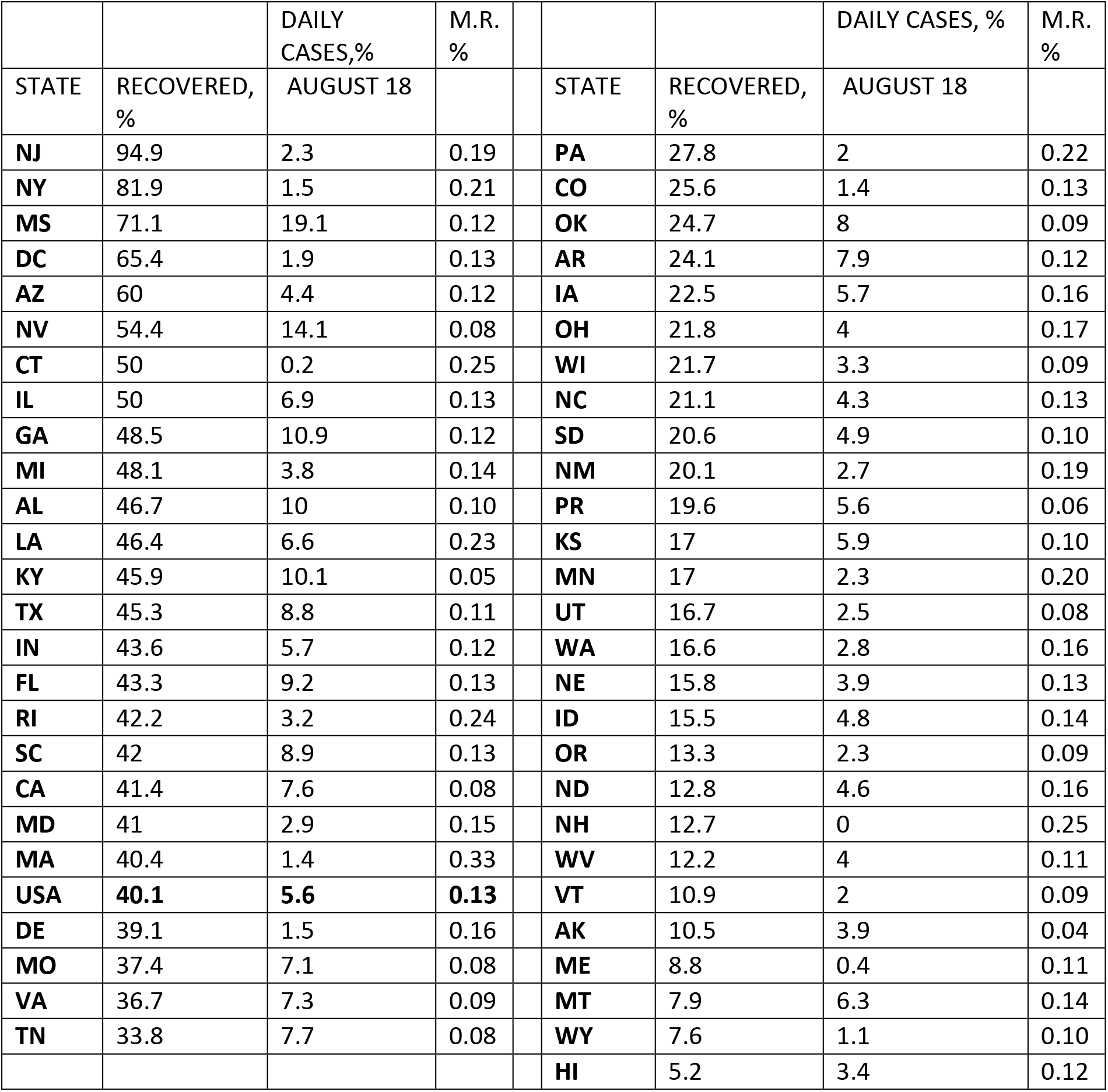
EPIDEMIC RECOVERED % POPULATION AND MORTALITY RATE (M.R.) BY STATE ON AUGUST 18, 2020.

The % Recovered column in Table 1 shows the population percent recovered from COVID-19. Vaccination program deployment would be most helpful in the states with the lower recovered population and higher incidence rates. Table 1 is a timetable, and the absolute numbers will shift with the advance of the epidemic while following the observed tendency.

## DISCUSSION

Over the short-term horizon, antibody testing and cumulated results of NHA cases are comparable. Rosenberg et al. conducted an antibody testing survey which showed a cumulative incidence of COVID-19 of 14% in New York state by March 29 (11). Silverman et al., using the CDC influenza surveillance networks to estimate the prevalence of SARS-CoV-2, found that over 8.3% of New York state residents were infected by March 28, while F.P. Havers et al. estimated the prevalence of SARS-CoV-2 at 6.9% over the period March 23 to April 1 (10, 22). This paper estimates the cumulative incidence of 9.6% on March 29. The infection rate was moving with a doubling time of about four days in New York state in March. Thus, slight differences in timing would lead to substantial differences in prevalence estimates. F.P. Havers et al. also performed a serological convenience survey in South Florida, showing a prevalence of 1.85% between March 23 and April 1, and 4.9% between April 6 and April 10. The present paper has estimates for the entire state at 0.65% and 2.0%, respectively.

Nevertheless, over the longer term, the longitudinal studies summarized here support the thesis that antibody levels fail to cumulate proportionately despite sustained rates of infections in the population. This decline is expected when the continuous production rate of antibodies in recovered patients decreases or ceases altogether since the half-life of IgA and IgM is 5 to 6 days, and for IgG 21 days (see Figure 5) (3,23).

To this point, the discovery of transient antibody response to SARS-CoV-2 has cast doubt on the value of subsequent serological studies aimed at determining the cumulative long-term prevalence of the virus. Additionally, the antibody response is graduated to the severity of the disease such that milder infections will lead to an earlier drop of detectable SARS-CoV-2 antibody below the baseline. Fan Wu et al. reported that 30% of cases tested had extremely low neutralizing antibodies specific for SARS-CoV-2, and another 17% had low levels (3). J Seow showed that patients with early modest antibody responses led to undetectable levels 50 days post-infection. Higher antibody responses in other individuals remain stable at 60 days, possibly longer (2). These findings suggest that current serological studies are limited to time-sliced views of the whole infected population and are more effective in detecting severe disease. Since severe disease is present only in a small subset of the afflicted population, results will tend to underestimate the population of recovered COVID-19 patients (1, 2).

Starting on week 21 of 2020, % positive PCR and new hospital admissions display close linear agreement with a coefficient of determination of 0.94 (see Figure 3A). This observation, together with the lower positive % PCR results, and non-decline of detected % positives with increasing test numbers, justify the association of % PCR positive cases with NHA. This close agreement forms the basis for estimating new cases between calendar weeks 10 and 20, when %PCR results show a positive bias.

Even in New Jersey, where according to this study, the recovered population has reached 95%, new cases continue to test daily at about 2% by PCR. This observation supports the notion that SARS-CoV-2 infection will continue to advance through the population, even when the recovered population’s extent is high. However, under this circumstance, the advance should happen at a substantially lower daily rate and, therefore, pose a lower immediate risk of contracting the infection to the remaining susceptible population, or overwhelming healthcare facilities and their human resources.

Postponing exposure makes sense because medical treatments demonstrated by the emergency use authorizations for Redemsivir (August 28), Dexamethasone (August 23) (24), convalescent plasma (August 23), and imminently promising treatments such as Calcifediol (25, 26, 27, 28), are becoming more effective over time.

The risk is highest for the over 65 age group, who account for 80% of all COVID-19 deaths in the United States while comprising 16% of the United States population. Within this age group, COVID-19 mortality is 9.5% of all deaths. By contrast, the age group below 24 has a 0.2% COVID-19 mortality rate, accounting for 0.9% of all deaths within the group, while comprising 32% of the United States (29).

Ultimately, a safe and effective vaccination will further reduce the risk if approved in a timely fashion. At the current daily rate of infection, 5% of the population, in 4 months, the recovered United States population would be at 66%.

Optimal deployment of a vaccine in limited supply should grant priority to the more susceptible states experiencing the higher daily levels of infection. This estimation technique is adaptable to the county level nationwide, provided the COVID-19 hospitalization rate for the locale is known. Ideally, this approach could be further informed by SARS-CoV-2 reactive T-cell prospective testing in potential vaccine subjects (30,31) given the existing cross-reactivity to SARS-CoV-2 and the limited ability antibody testing displays in discerning between naïve and recovered populations.

More effective mitigation measures should focus on safeguarding the population above 65 years of age to reduce mortality substantially and efficiently (14). These measures should emphasize distancing, isolation, technological medical monitoring and connections, and assure that the elderly are receiving the correct nutritional supplementation, including monitoring their vitamin D levels to ensure blood levels are at or above 50 ng/ml (25,26,27,28).

An additional utility of relying on new hospitalizations instead of PCR is that since hospitalization rate is independent of testing, it should provide a more accurate assessment of the daily infection rate when testing is reduced so apparent % infected increase. For instance, on August 18, the NHA % positive cases in Arizona are 4.4%, while the PCR dependent test yields 16.3%; Florida shows 9.3% for NHA cases versus 14.7% for PCR; South Dakota comes to 4.1% compared to 11.5% for PCR. For states that continued testing at high numbers such as New York, the NHA % positive cases are 1.5% while the PCR is 1.0%.

## METHODS

### Estimation of daily % Infected Population based on New Hospital Admissions

Daily % Infected Population based on hospital admissions is described by the expression: %NHA cases = Observed daily NHA/(Constant Ratio NHA/%PCR)

Constant Ratio NHA/ %PCR = Average NHA (June 25 to August 31)/Average % PCR (June 25 to August 31) or 516

### Estimation of New Hospital Admissions

The New Hospital Admissions daily value is computed from the reported ‘hospitalized currently’ data column by assuming a 12-day average hospital stay, which is applied to create a daily hospital discharge value (1, 8, 12) described by the expressions:

> NHA = New net hospitalizations + Hospital discharge

New net hospitalizations = hospitalizations current (today)-hospitalizations current (yesterday) Hospital discharge = hospitalizations current (today) / 12

Florida and Kansas did not report current hospitalizations over a sufficient period to compute NHA for this analysis. Instead, both reported cumulative hospitalizations. In both cases, NHA was calculated from this data by subtracting today’s cumulative hospitalizations from the previous day. Other entities included in the COVID-19 database (G.U., MP, and VI) were excluded from the analysis because of data deficiencies.

For 10 states, cumulative hospitalizations and currently hospitalized are reported for the United States in the COVID-19 tracking database from The Atlantic (1). The resulting comparison (see Table Cumulative versus Currently hospitalized) supports the choice of 12 days as the median time for hospital discharge.

The second column of Table 1 shows the ratio obtained for new hospital admissions when computed by subtracting following day cumulative headcount versus the outcome of the algorithm described above derived from currently hospitalized data when the discharge rate is set at 12 days.

An alternate way of calculating this ratio is presented in the third column, which calculates the hospital stay number of days for each state ratio to match the ‘currently hospitalized’ algorithm to the cumulative calculation. The median for each approach is 11.4 days, while the combined average value is 12.7 days. The 95% confidence interval of the mean for each is about 25%.

### Recovered population estimation

Knowing the time that PCR tests can detect the virus during the disease timeline is essential to estimate the cumulative recovered population from daily % positive cases. Literature estimates for the median time to conversion to PCR negative range from 17 to 28 days (14, 17, 18, 19, 20, 21).

It is possible to assess this range’s feasibility by calculating % recovered population obtained from these values. The extent of the calculated % recovered population varies inversely with the PCR detection length. Postulating that no state today is above 100%, the detection length cannot be less than 26.6 days. It is impossible to deduce a higher boundary, but the literature does not support a number above 28 days, which justifies the selection of 28 days for the PCR detection period (9,13,14,15,16,17,18,19,20,21,32,33).

A separate line of evidence based on anecdotal medical experience with COVID-19 also supports a range for viral shedding median exceeding 19 days. Average incubation time is reported at 4 to 5 days, and reportedly, patients become PCR positive 1 to 2 days before symptoms onset. From the emergence of symptoms to dyspnea, the median time-lapse is about seven days. The median length of hospital stays come to 12 days, after which patients are PCR negative. Adding these consecutive periods together returns a positive PCR period of at least 20 days (13,14,15,16,33,34,35).

After correcting the daily percent positive PCR results for sampling bias via the number of hospital admissions yields daily % COVID-19 positive cases. This number divided by 28 days of viral detectability returns the daily recovered population % of the pandemic. Sequentially summing this daily estimate through the time series results in the cumulative recovered population.

### Comparison of SARS-CoV-2 Antibodies longitudinal time course versus cumulation of recovered COVID-19 cases

SARS-CoV-2 antibodies fail to cumulate effectively in the recovered population falling below the detectability level over time (2,3). This observation is explainable by the observed transient response of antibody synthesis triggered by the infection. In such cases, because of the limited half-life of IgA (5 days), igM (6 days), and IgG (21 days), the antibody presence declines past its production peak with a tendency toward a steady-state concentration, or disappearance depending on whether the reduced remaining level of antibody production exceeds removal (23).

The COVID-19 database published by the Atlantic captures the antibody tests time course for four states: Colorado, Georgia, South Carolina, and Texas (see Figure 5, a, b, c, and d, respectively). In Figure 5a, the antibody population % value for Colorado remains unchanged from May 9 to September 6, even though during this time, the daily infected population averages 2.8%. The neutral slope suggests that the rate of new antibody production at this infection level matches the decay rate. Figure 5b shows that starting on July 18, the antibody population % for Georgia increases by 5.4% compared to the observed daily infection average rate of 13.1%. Similar behavior is apparent in South Carolina and Texas, where antibody cumulation fails to account for the daily infection rate (see Figure 5c and 5d, respectively).

### Limitations

The ratio of New Hospital Admissions (NHA) to %PCR positives is used to calculate the percentage of the population infected when the total daily PCR test numbers are below 500.000 from March 18 to June 20. The ratio was calculated from the data reported from June 21 to August 31. The average value for the ratio over this time when daily tests exceeded 500.000 is 516 NHA/%PCR cases in the United States, and its 95% confidence interval is 10.5 units. As the ratio’s value decreases, the % recovered increases approaching the value obtained from % PCR positives without mitigating for testing number bias. This calculation’s validity rests on the accuracy of the data reported in the COVID Tracking Project at the Atlantic (1).

The duration of PCR viral detection in infected people affects the estimation of the nationwide prevalence of the infection proportionately. Multiple studies support the conservative choice of 28 days for the PCR detectability period. However, such studies are limited to less than 300 patients and span a limited calendar time window. The selected 28-day value sets an upper limit for the % recovered population estimated here. Similarly, the disease severity, medical treatments, and fragility of the remaining population could influence PCR detectability length. If these tend to reduce the detectability window as the epidemic advances, the estimates presented here will underestimate the recovered population.

Likewise, the length of hospitalization set at 12 days is consistent with that found in the ten states reporting the information directly. However, individual states’ reported data is more susceptible to variability leading to the 95% confidence interval of the mean at 24 to 27% (3 days). This variability also affects the daily case % estimated from NHA and, consequentially, the % recovered population.

## Data Availability

All data is available

https://covidtracking.com/

